# Functional Impairment, Pain, and Fatigue Among Long COVID Survivors in Bangladesh: A Multisite Cross-Sectional Study

**DOI:** 10.64898/2026.09.21.26363589

**Authors:** Polok Halder, Md. Asadul Islam, Suprantha Sutra Dhar, Shahnur Zinia Parvin, Fyzul Kabir, Md. Shohag Rana, Farhin Islam Peya, Md. Nesaruddin

**Author notes:** Corresponding author: Polok Halder, Clinical Physiotherapist, BRB Hospitals Limited, 77/A Panthapath, Dhaka-1215, Bangladesh.

## Abstract

**Background:** Long COVID can involve persistent symptoms, functional limitations, pain, and fatigue. Evidence describing these inter-related outcomes in Bangladeshi survivors remains limited. This study examined functional status and factors associated with pain and fatigue among long COVID survivors in Bangladesh.

**Methods:** A multisite cross-sectional study included 240 adults with persistent symptoms after COVID-19 who were recruited from five healthcare facilities in Bangladesh using convenience sampling. Data were collected using a structured questionnaire, the Post-COVID-19 Functional Status scale, the Brief Pain Inventory, and the Brief Fatigue Inventory. Adjusted ordinal logistic regression models were used for functional-status domains, multiple linear regression models were used for pain and fatigue outcomes, and Spearman rank correlation analysis assessed relationships among continuous measures.

**Results:** The mean age was 50.3 years (SD 14.5), and 176 participants (73.3%) were female. Slight limitation was most common for constant care (138 [57.5%]), instrumental activities of daily living (120 [50.0%]), and social roles (125 [52.1%]), whereas moderate limitation predominated for basic activities of daily living (149 [62.1%]). Cough (177 [73.8%]), lack of appetite (170 [70.8%]), chest pain (161 [67.1%]), and pain (155 [64.6%]) were the most frequently reported persistent symptoms. Older age was associated with greater ADL impairment (aOR 1.02, 95% CI 1.00–1.05; Wald χ^2^=3.95, p=0.047). In adjusted linear models, booster vaccination was associated with lower pain and fatigue scores across all reported BPI and BFI outcomes, while tobacco non-use was associated with lower scores across several domains. Pain and fatigue measures showed strong positive correlations.

**Conclusions:** Long COVID survivors in this Bangladeshi multisite cohort experienced substantial functional limitations together with prominent pain and fatigue. The observed associations with vaccination status, tobacco use, sociodemographic factors, and physiotherapy exposure should be interpreted as observational associations rather than causal effects. Prospective studies are needed to clarify determinants of recovery and the effectiveness of rehabilitation strategies.

**Preprint status:** This manuscript is a preliminary research report that has not undergone peer review.

## Introduction

Long COVID, also referred to as post-COVID-19 condition, encompasses persistent or new symptoms following acute SARS-CoV-2 infection. The World Health Organization defines post-COVID-19 condition as symptoms occurring in individuals with a history of probable or confirmed infection, usually three months from onset, lasting for at least two months and not explained by an alternative diagnosis [1]. Studies have documented prolonged fatigue, dyspnea, pain, cognitive complaints, and other multisystem manifestations that can continue after acute infection and interfere with daily activities and social participation [2-5].

Functional recovery is an important dimension of long-term COVID-19 outcomes. The Post-COVID-19 Functional Status scale was developed to capture the consequences of persistent symptoms on everyday activities and participation [6]. Pain and fatigue are also clinically important because they can contribute to functional limitations and may occur together. The Brief Pain Inventory and Brief Fatigue Inventory provide structured measures of symptom severity and interference [7,8].

Rehabilitation, including physiotherapy, has been proposed as one component of multidisciplinary long COVID management. Previous reports describe respiratory, musculoskeletal, exercise-based, and individualized rehabilitation approaches for persistent symptoms [9-12]. However, observational evidence from Bangladesh remains limited, particularly regarding the relationships among functional impairment, pain, fatigue, vaccination status, tobacco exposure, and rehabilitation exposure.

The present study examined functional limitations and symptom burden among Bangladeshi long COVID survivors and evaluated demographic, clinical, vaccination, tobacco, and physiotherapy-related factors associated with functional status, pain, and fatigue.

## Materials and Methods

### Study design and setting

This was a multisite cross-sectional study conducted among adults with persistent symptoms following COVID-19. Participants were recruited from Enam Medical College and Hospital, Z H Sikder Women’s Medical College and Hospital, KC Hospital and Diagnostic Center Ltd., Shin Shin Japan Hospital, and Nostrum Hospital in Bangladesh.

### Participants and sampling

Participants were adults who had experienced COVID-19 and reported persistent post-COVID symptoms. Convenience sampling was used, and 240 participants were included. Eligibility criteria included adult age, documented COVID-19 infection, persistent long COVID symptoms, and provision of informed consent. Participants who were medically unstable, unable to provide informed consent, or otherwise met the study exclusion criteria were excluded.

### Measures

The BPI and BFI are copyrighted instruments. The instruments were used for research assessment in accordance with the applicable source and permission requirements.

For clarity, COVID-negative status refers to a negative COVID-19 test recorded at assessment and does not indicate absence of a previous COVID-19 infection.

### Statistical analysis

Continuous variables are presented as means with standard deviations, and categorical variables as frequencies and percentages. Adjusted ordinal logistic regression models were fitted separately for the reported functional-status domains. The models adjusted for age, gender, marital status, education, occupation, living area, income, vaccination status, COVID-negative status, hospitalisation, ICU admission, hospital stay duration, tobacco use, treatment type, physiotherapy receipt, type of physiotherapy, and physiotherapy provider. For ordinal logistic regression, coefficient significance was assessed using Wald χ^2^ tests, with adjusted odds ratios (aORs) and 95% confidence intervals (CIs) reported.

Multiple linear regression models were used for the BPI and BFI outcomes with the same covariate set. Regression coefficients (β), 95% CIs, t statistics, and two-sided p-values are reported. Spearman rank correlation was used to examine relationships among age and pain/fatigue measures; Spearman’s ρ is reported as the correlation test statistic. A two-sided p-value <0.05 was considered statistically significant. Results with p-values between 0.05 and 0.10 were treated as borderline associations and are not presented as statistically significant.

### Ethical considerations

The study received ethical approval from the Institutional Review Board of Bangladesh Health Professions Institute (BHPI), Centre for the Rehabilitation of the Paralysed (CRP), Savar, Dhaka, Bangladesh (approval reference CRP-BHPI/IRB/10/2022/673). Ethical approval was granted at the 33rd IRB meeting held on September 24, 2022. Written informed consent was obtained from all participants. Institutional permission for data collection was obtained from the participating facilities.

## Results

### Participant characteristics

A total of 240 long COVID survivors were included. The mean age was 50.3 years (SD 14.5); 176 participants (73.3%) were female, 136 (56.7%) were married, and 153 (63.7%) lived in urban areas. Higher secondary education was reported by 73 participants (30.4%), and 95 (39.6%) were in job/services occupations. Two vaccine doses had been received by 169 participants (70.4%), while 17 (7.1%) had received a booster. A total of 136 participants (56.7%) had been hospitalised, 136 (56.7%) reported ICU admission, 127 (52.9%) reported tobacco use, and 134 (55.8%) reported receiving physiotherapy after recovery. Detailed characteristics are presented in Table 1.

**Table 1.** Participant characteristics (N=240)

| Variable | n (%) or mean (SD) |
| --- | --- |
| Age, mean (SD) | 50.3 (14.5) |
| Age 20–30 | 51 (21.3) |
| Age 31–40 | 17 (7.1) |
| Age 41–50 | 45 (18.8) |
| Age 51–60 | 37 (15.4) |
| Age ≥60 | 90 (37.5) |
| Male | 64 (26.7) |
| Female | 176 (73.3) |
| Married | 136 (56.7) |
| Unmarried | 37 (15.4) |
| Widow | 27 (11.3) |
| Divorcee | 40 (16.7) |
| No formal education | 12 (5.0) |
| Primary | 60 (25.0) |
| Secondary | 55 (22.9) |
| Higher secondary | 73 (30.4) |
| Bachelor or above | 40 (16.7) |
| Job/Services | 95 (39.6) |
| Farmers | 15 (6.3) |
| Business | 28 (11.7) |
| Freelancer | 27 (11.3) |
| Others | 75 (31.3) |
| Rural | 39 (16.3) |
| Semi-urban | 48 (20.0) |
| Urban | 153 (63.7) |
| Income 0–15k BDT | 28 (11.7) |
| Income 16–30k BDT | 91 (37.9) |
| Income 31–45k BDT | 50 (20.8) |
| Income ≥46k BDT | 71 (29.6) |
| No COVID vaccination | 21 (8.8) |
| First dose | 33 (13.8) |
| Second dose | 169 (70.4) |
| Booster | 17 (7.1) |
| COVID-negative status | 142 (59.2) |
| Hospitalisation | 136 (56.7) |
| ICU admission | 136 (56.7) |
| Hospital stay 0–10 days | 169 (70.4) |
| 11–20 days | 61 (25.4) |
| ≥21 days | 10 (4.2) |
| Tobacco use | 127 (52.9) |
| Medicine only | 143 (59.6) |
| Medicine + oxygen | 62 (25.8) |
| Ventilation | 35 (14.6) |
| Received physiotherapy | 134 (55.8) |
| Respiratory physiotherapy | 52 (21.7) |
| Cardiovascular physiotherapy | 62 (25.8) |
| Musculoskeletal physiotherapy | 71 (29.6) |
| Neurological physiotherapy | 37 (15.4) |
| Other physiotherapy | 18 (7.5) |
| Physiotherapist | 65 (27.1) |
| Medical technologist | 57 (23.8) |
| Physician | 50 (20.8) |
| Social media | 30 (12.5) |
| YouTube | 14 (5.8) |
| Self | 24 (10.0) |

### Post-COVID functional status and symptom burden

Slight limitation was most frequent for constant care (138 [57.5%]), instrumental activities of daily living (120 [50.0%]), and participation in usual social roles (125 [52.1%]). Moderate limitation predominated for basic activities of daily living (149 [62.1%]). No participant reported no limitation for basic ADL, instrumental ADL, or social roles. Mean BPI pain severity was 19.03 (SD 3.20), and pain interference was 36.77 (SD 7.06). Mean BFI fatigue severity was 14.27 (SD 2.82), and fatigue interference was 31.24 (SD 5.78).

**Table 2.**
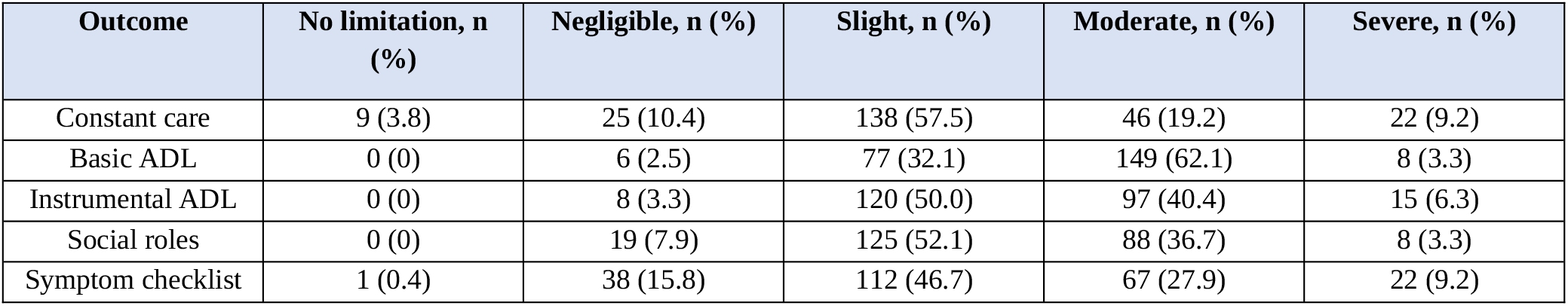
Post-COVID functional status domains (N=240)

| Outcome | No limitation, n (%) | Negligible, n (%) | Slight, n (%) | Moderate, n (%) | Severe, n (%) |
| --- | --- | --- | --- | --- | --- |
| Constant care | 9 (3.8) | 25 (10.4) | 138 (57.5) | 46 (19.2) | 22 (9.2) |
| Basic ADL | 0 (0) | 6 (2.5) | 77 (32.1) | 149 (62.1) | 8 (3.3) |
| Instrumental ADL | 0 (0) | 8 (3.3) | 120 (50.0) | 97 (40.4) | 15 (6.3) |
| Social roles | 0 (0) | 19 (7.9) | 125 (52.1) | 88 (36.7) | 8 (3.3) |
| Symptom checklist | 1 (0.4) | 38 (15.8) | 112 (46.7) | 67 (27.9) | 22 (9.2) |

BPI and BFI scores: pain severity 19.03 (3.20); pain interference 36.77 (7.06); pain affective interference 15.52 (3.70); pain physical interference 21.25 (5.12); fatigue severity 14.27 (2.82); fatigue interference 31.24 (5.78); fatigue affective interference 15.60 (2.85); and fatigue physical interference 15.63 (4.37).

The eight most common persistent symptoms were cough (177 [73.8%]), lack of appetite (170 [70.8%]), chest pain (161 [67.1%]), pain (155 [64.6%]), dyspnoea (139 [57.9%]), fatigue (120 [50.0%]), anosmia (119 [49.6%]), and headache (117 [48.8%]).

**Table 3.**
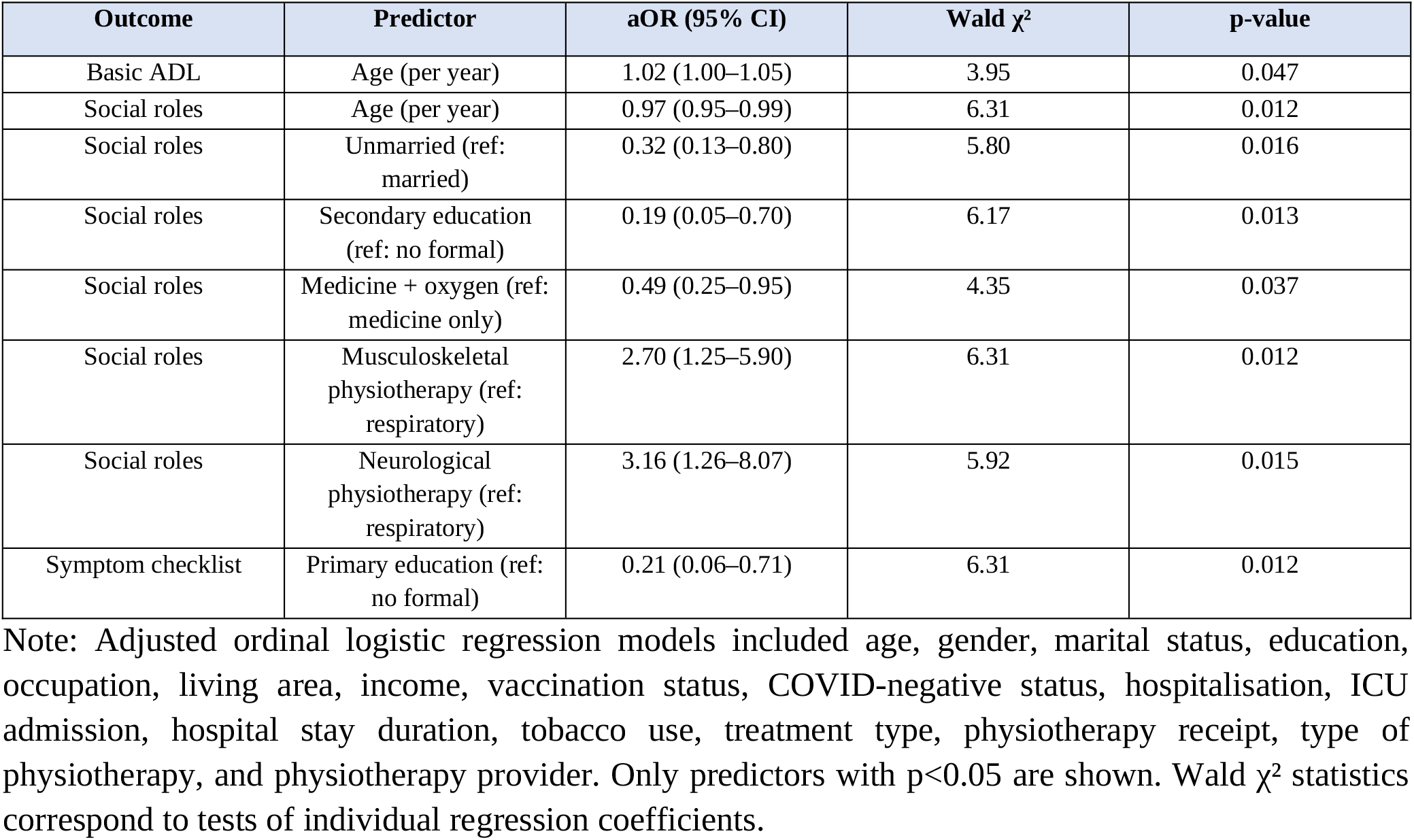
Ordinal regression: significant predictors of functional-status domains.

### Factors associated with functional impairment

Adjusted ordinal logistic regression models were fitted for each functional-status domain. Older age was associated with greater ADL impairment (aOR 1.02, 95% CI 1.00–1.05; Wald χ^2^=3.95, p=0.047). For social-role participation, older age (aOR 0.97, 95% CI 0.95–0.99; Wald χ^2^=6.31, p=0.012), unmarried status (aOR 0.32, 95% CI 0.13–0.80; Wald χ^2^=5.80, p=0.016), secondary education (aOR 0.19, 95% CI 0.05–0.70; Wald χ^2^=6.17, p=0.013), and medicine plus oxygen treatment (aOR 0.49, 95% CI 0.25–0.95; Wald χ^2^=4.35, p=0.037) were associated with lower odds of worse social-role impairment. Musculoskeletal physiotherapy (aOR 2.70, 95% CI 1.25–5.90; Wald χ^2^=6.31, p=0.012) and neurological physiotherapy (aOR 3.16, 95% CI 1.26–8.07; Wald χ^2^=5.92, p=0.015) were associated with higher odds of worse social-role impairment. Primary education was associated with lower odds of worse symptom-checklist status (aOR 0.21, 95% CI 0.06–0.71; Wald χ^2^=6.31, p=0.012). No predictor reached p<0.05 for constant care or instrumental ADL.

### Factors associated with pain and fatigue

Multiple linear regression models identified several associations with BPI and BFI outcomes. Business occupation was associated with higher pain severity (β=+2.20, 95% CI 0.87–3.53; t=3.34, p=0.001). Not being admitted to ICU was associated with lower pain severity (β=−0.96, 95% CI −1.83 to −0.09; t=−2.16, p=0.032), as was tobacco non-use (β=−1.20, 95% CI −2.04 to −0.35; t=−2.78, p=0.006).

Booster vaccination was associated with lower pain severity (β=−3.22, 95% CI −5.52 to −0.92; t=−2.73, p=0.007).

For pain interference, non-hospitalisation (β=−2.46, 95% CI −4.58 to −0.34; t=−2.27, p=0.024), tobacco non-use (β=−2.34, 95% CI −4.12 to −0.56; t=−2.57, p=0.011), and booster vaccination (β=−10.01, 95% CI −14.81 to −5.21; t<−3.34, p<0.001) were associated with lower scores. For pain affective interference, occupation in the ‘other’ category (β=+1.18, 95% CI 0.13–2.22; t=2.20, p=0.029) and semi-urban residence (β=+1.55, 95% CI 0.06–3.04; t=2.05, p=0.042) were associated with higher scores, whereas non-receipt of physiotherapy (β=−1.16, 95% CI −2.11 to −0.21; t=−2.41, p=0.017) and booster vaccination (β=−4.42, 95% CI −6.93 to −1.91; t<−3.34, p<0.001) were associated with lower scores. For pain physical interference, widowhood was associated with higher scores (β=+2.52, 95% CI 0.15–4.90; t=2.09, p=0.038), while tobacco non-use (β=−1.87, 95% CI −3.25 to −0.50; t=−2.68, p=0.008) and booster vaccination (β=−5.58, 95% CI −9.31 to −1.85; t=−2.73, p=0.004) were associated with lower scores.

For fatigue severity, unmarried status (β=+1.22, 95% CI 0.06–2.38; t=2.06, p=0.041) and business occupation (β=+1.45, 95% CI 0.33–2.57; t=2.54, p=0.012) were associated with higher scores, whereas non-receipt of physiotherapy (β=−0.89, 95% CI −1.62 to −0.15; t=−2.37, p=0.019) and booster vaccination (β=−4.09, 95% CI −6.04 to −2.14; t<−3.34, p<0.001) were associated with lower scores. Tobacco non-use and booster vaccination were associated with lower fatigue interference (β=−2.10, 95% CI −3.61 to −0.58; t=−2.73, p=0.007; and β=−7.07, 95% CI −11.17 to −2.97; t<−3.34, p<0.001, respectively). For fatigue affective interference, tobacco non-use (β=−0.96, 95% CI −1.71 to −0.20; t=−2.51, p=0.013) and booster vaccination (β=−3.33, 95% CI −5.36 to −1.29; t=−3.13, p=0.002) were associated with lower scores. Widowhood was associated with higher fatigue physical interference (β=+2.27, 95% CI 0.20–4.35; t=2.16, p=0.032), whereas booster vaccination was associated with lower scores (β=−3.74, 95% CI −6.99 to −0.49; t=−2.26, p=0.025).

**Table 4.**
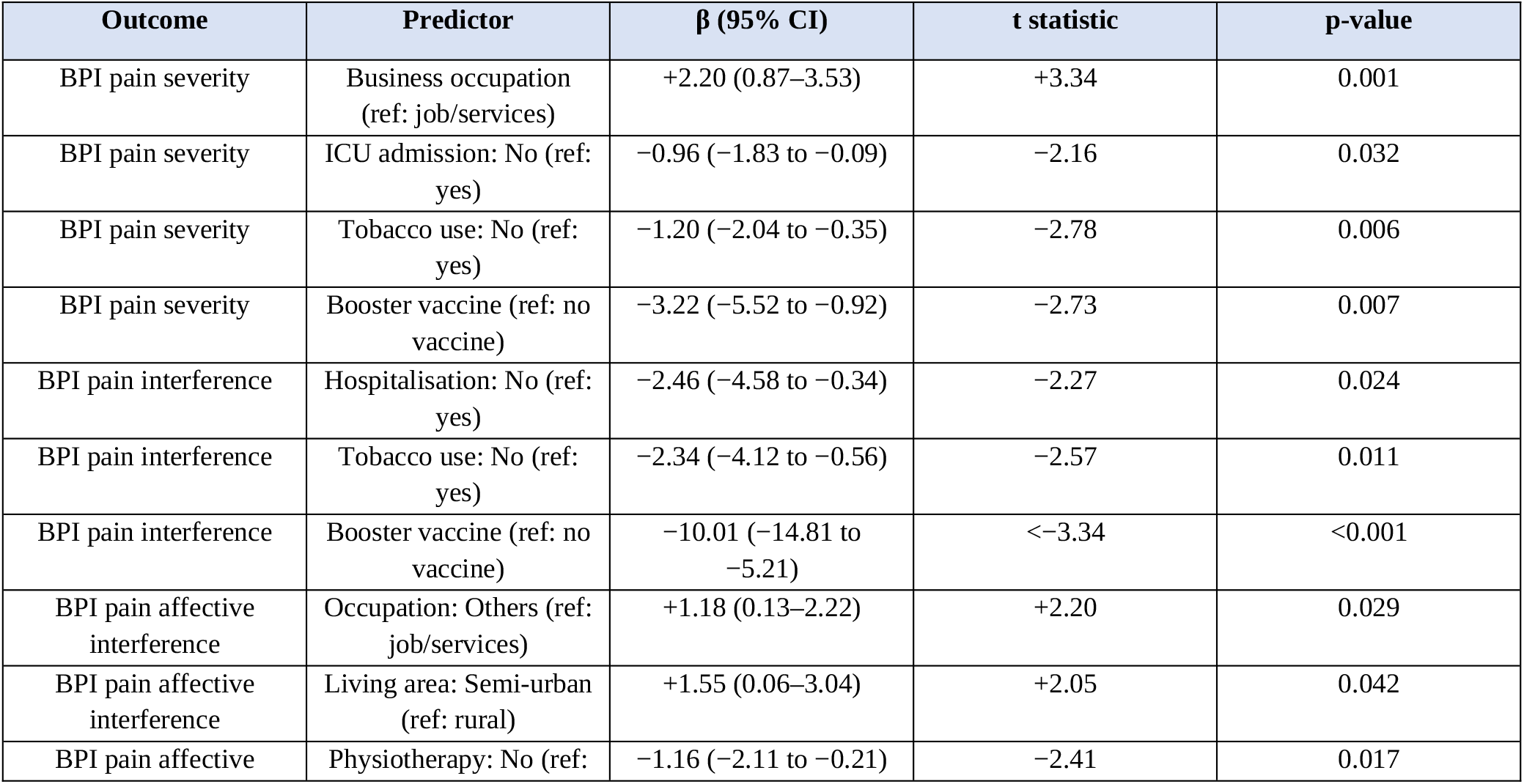
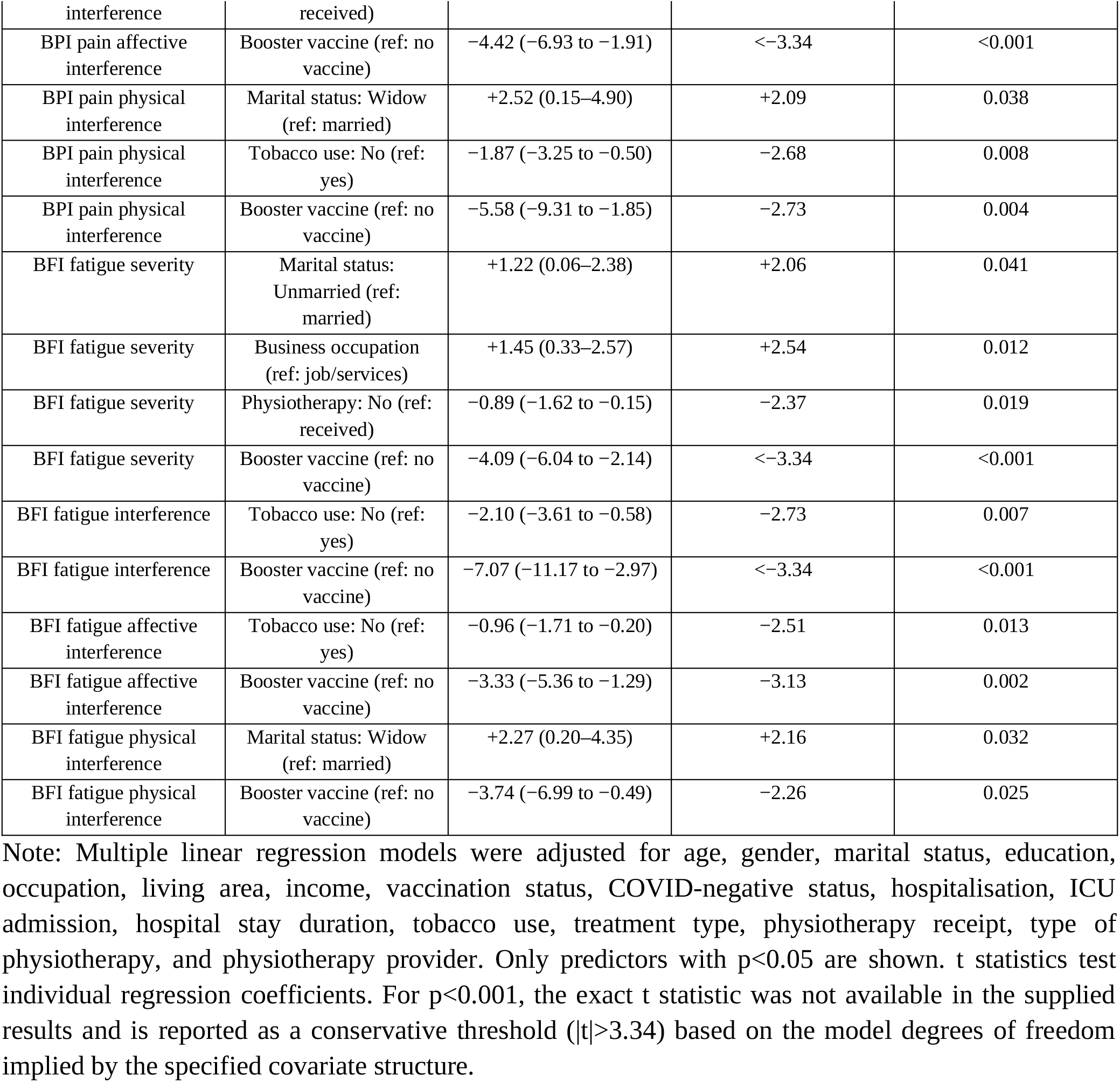
Linear regression: significant predictors of BPI and BFI scores.

### Correlation analysis

Spearman correlation analysis showed strong positive relationships among pain severity, pain interference, fatigue severity, and fatigue interference. Pain severity correlated strongly with pain interference (ρ=0.85, p<0.001), fatigue severity (ρ=0.78, p<0.001), and fatigue interference (ρ=0.82, p<0.001). Age showed weak correlations with most outcomes (|ρ|<0.20). These findings indicate that pain and fatigue were closely inter-related in this cohort.

## Discussion

This multisite study describes a substantial burden of functional limitation, pain, and fatigue among 240 long COVID survivors in Bangladesh. Moderate limitation was particularly common in basic activities of daily living, while slight limitation predominated in several other functional domains. The strong correlations among pain and fatigue measures suggest that these symptoms may represent interconnected components of the post-COVID symptom burden rather than isolated complaints.

Older age was associated with greater impairment in basic activities of daily living. This association may reflect the interaction between age-related reductions in physiological reserve and persistent postinfectious symptoms. The associations observed for social-role participation should be interpreted cautiously because cross-sectional data cannot establish temporality and may be influenced by baseline characteristics, social circumstances, healthcare access, and residual confounding.

Booster vaccination was associated with lower scores across all BPI and BFI outcomes reported in the analysis. The consistency of these associations is notable, but the findings should not be interpreted as evidence that booster vaccination caused lower symptom burden. Only 17 participants (7.1%) had received a booster, and vaccination status may be related to age, healthcare access, health behavior, timing of infection, and other unmeasured factors. The association should therefore be considered hypothesis-generating and evaluated in larger longitudinal studies.

Tobacco non-use was associated with lower scores across several pain and fatigue domains. This pattern is compatible with a possible relationship between tobacco exposure and persistent symptom burden, but the present cross-sectional design does not permit causal inference and residual confounding cannot be excluded.

The associations involving physiotherapy require particular caution. Musculoskeletal and neurological physiotherapy were associated with higher odds of worse social-role impairment, while non-receipt of physiotherapy was associated with lower scores for selected pain and fatigue outcomes. These findings should not be interpreted as evidence that physiotherapy worsened outcomes. Confounding by indication is a plausible explanation because participants with greater functional impairment or more complex symptoms may have been more likely to receive or be referred for rehabilitation. Differences in timing, type, intensity, and duration of rehabilitation may also have contributed.

The findings have implications for rehabilitation assessment. Long COVID care should consider functional status, symptom severity, participation, pain, and fatigue together rather than focusing on a single symptom. Individualized rehabilitation planning may need to account for symptom clustering, severity of the acute illness, tobacco exposure, and social participation. Recent rehabilitation trials and reviews support continued investigation of structured and multidisciplinary approaches, while also highlighting the need for appropriately designed prospective studies [13-16].

Overall, the findings support a multidimensional approach to long COVID assessment. Longitudinal studies with larger and more representative samples are needed to determine symptom trajectories, identify modifiable factors, and evaluate the effectiveness and timing of individualized rehabilitation interventions.

## Limitations

This study has several limitations. First, the cross-sectional design prevents causal inference and does not allow assessment of symptom trajectories over time. Second, convenience sampling from five healthcare facilities may limit generalizability to the wider Bangladeshi population. Third, the booster-vaccinated subgroup was small, limiting the precision of estimates for this comparison. Fourth, several variables were self-reported and may be affected by recall or reporting bias. Fifth, the study included multiple outcomes and predictors, increasing the possibility of chance findings. Finally, physiotherapy exposure was not assigned prospectively, and treatment timing, intensity, duration, and modality were not standardized; consequently, the observed rehabilitation associations cannot be interpreted as treatment-effect estimates.

## Conclusions

Bangladeshi long COVID survivors in this multisite cohort experienced substantial functional limitations accompanied by considerable pain and fatigue. Pain and fatigue were strongly inter-related, while age, sociodemographic characteristics, tobacco exposure, vaccination status, and rehabilitation exposure showed associations with selected functional and symptom outcomes. The consistent association between booster vaccination and lower pain and fatigue scores warrants prospective investigation, particularly in larger and more representative populations.

Future research should use longitudinal designs and standardized rehabilitation protocols to clarify determinants of recovery and evaluate the effectiveness, timing, and type of physiotherapy interventions for long COVID. Clinical assessment should remain individualized and should consider functional status, symptom burden, and participation together.

## Supporting information

Supplementary Statistical Tables

STROBE Checklist

## Data Availability

The data supporting the findings are not publicly available because they contain participant-level research information. Data may be made available from the corresponding author on reasonable request and subject to applicable ethical and institutional restrictions.

## Declarations

### Ethics approval

The study was approved by the Institutional Review Board of the Bangladesh Health Professions Institute (BHPI), Centre for the Rehabilitation of the Paralysed (CRP), Savar, Dhaka, Bangladesh (approval reference CRP-BHPI/IRB/10/2022/673). Ethical approval was granted at the 33rd IRB meeting held on September 24, 2022.

### Consent to participate

Written informed consent was obtained from all participants before participation in the study.

### Conflict of interest

The authors declare that they have no competing interests related to this study or its publication.

### Funding

The authors received no specific external funding for this study.

### Data availability

The data generated and analyzed during the current study are not publicly available because they contain participant-level information and are subject to ethical and institutional restrictions. Data may be made available by the corresponding author upon reasonable request, subject to approval by the relevant institutional and ethical authorities.

## Author contributions

Dr. Md. Nesaruddin: Conceptualization, methodology, data collection, investigation, and critical revision of the manuscript.

Dr. Farhin Islam Peya: Methodology, data collection, investigation, and critical revision of the manuscript.

Dr. Md. Shohag Rana: Methodology, investigation, data collection, and critical revision of the manuscript.

Dr. Fyzul Kabir: Methodology, investigation, and critical revision of the manuscript.

Dr. Shahnur Zinia Parvin: Data collection, investigation, and critical revision of the manuscript.

Dr. Suprantha Sutra Dhar: Methodology, supervision, investigation, and critical revision of the manuscript.

Dr. Md. Asadul Islam: Data collection, investigation, interpretation of findings, and critical revision of the manuscript.

Dr. Polok Halder: Conceptualization, methodology, data curation, formal analysis, interpretation of findings, manuscript drafting, and critical revision of the manuscript.

All authors reviewed and approved the final version of the manuscript and agreed to be accountable for all aspects of the work.

## Acknowledgments

The authors acknowledge the Bangladesh Health Professions Institute and the participating healthcare facilities for supporting the study and the participants who contributed their time and information.

## Figure legends

**Figure 1.**
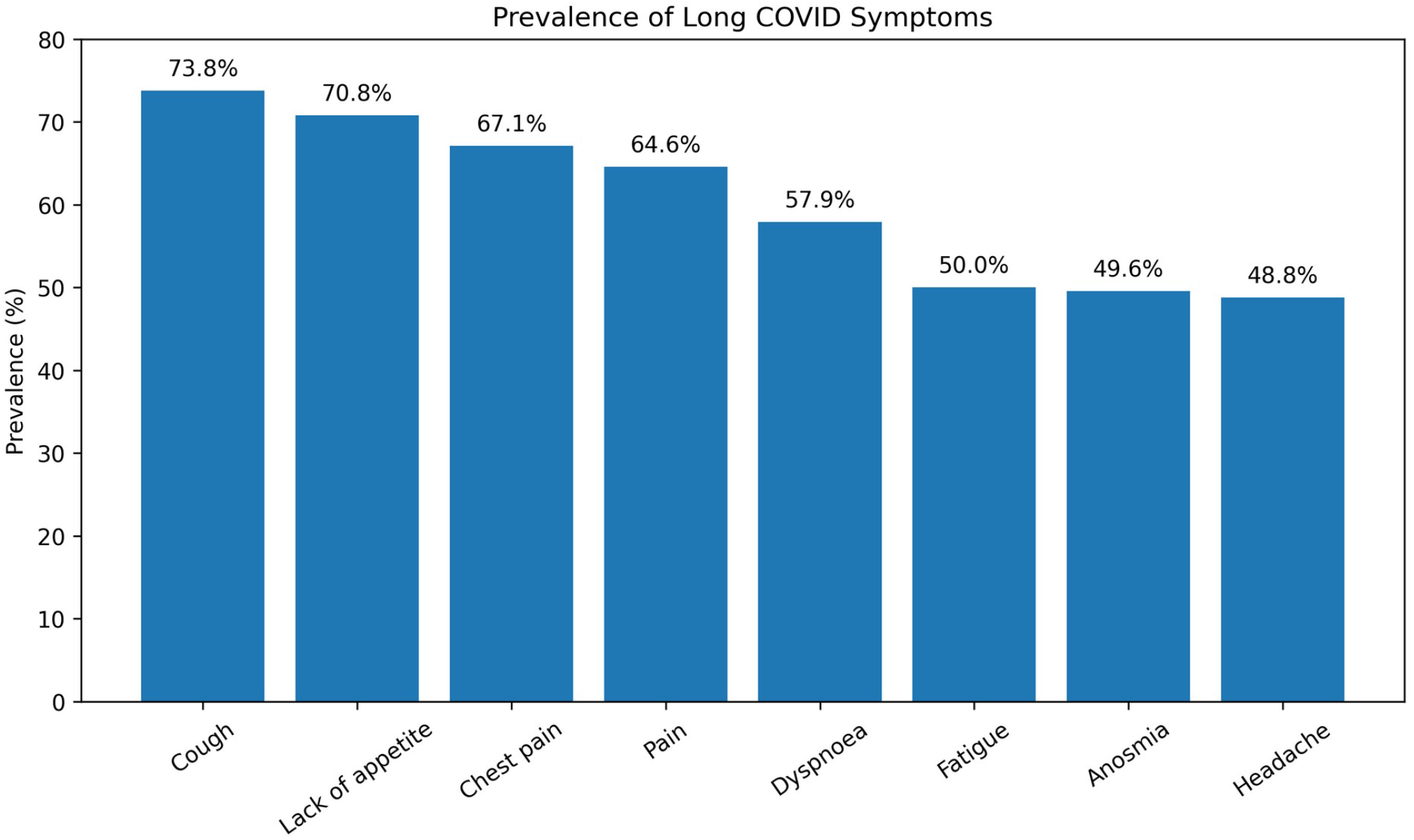
Prevalence of long COVID symptoms. Bar chart showing the percentage of participants (N=240) reporting the eight most common persistent symptoms.

**Figure 2.**
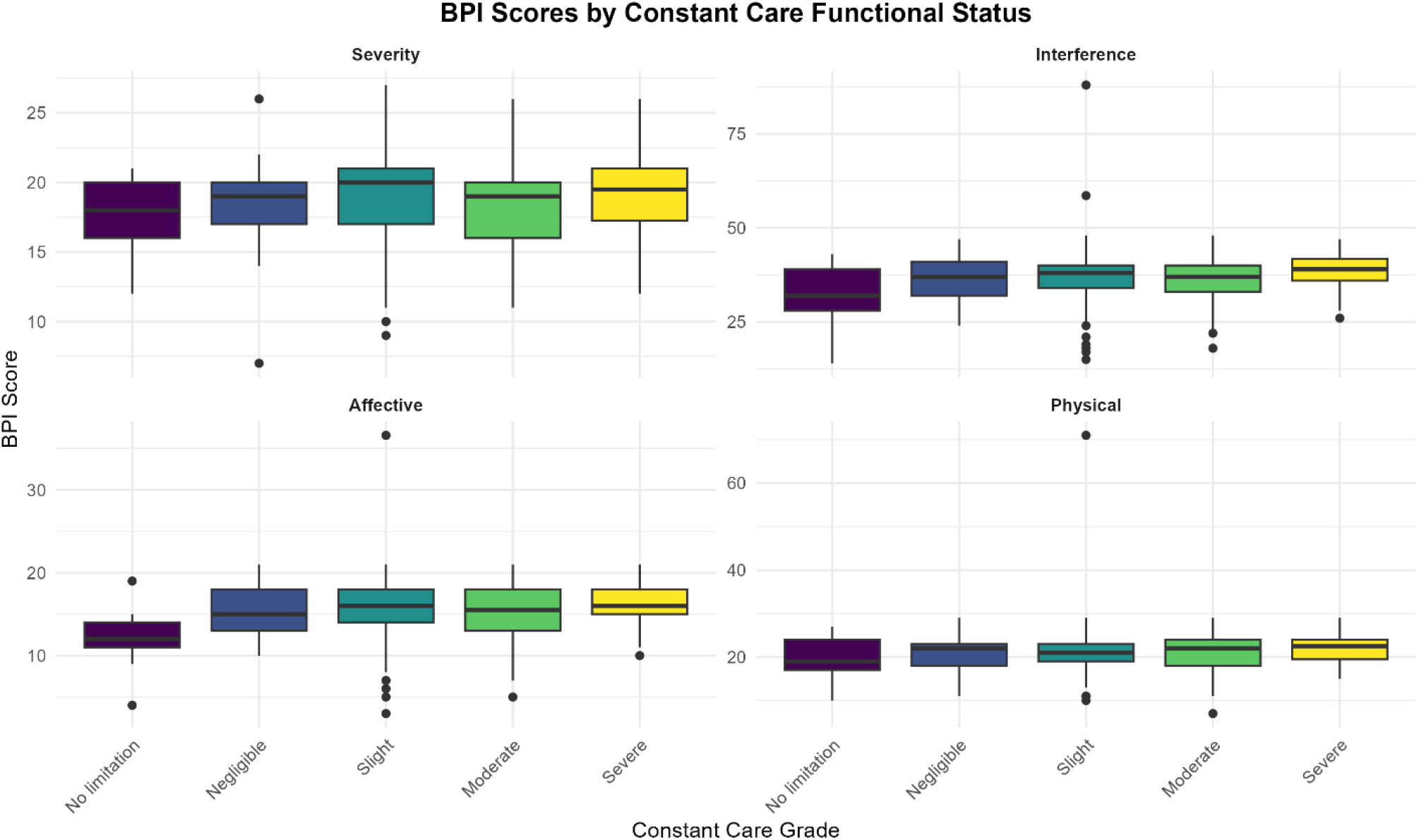
Boxplots of BPI scores by constant care functional status. Distribution of Brief Pain Inventory subscale scores across the five grades of the constant care domain of the Post-COVID-19 Functional Status scale.

**Figure 3.**
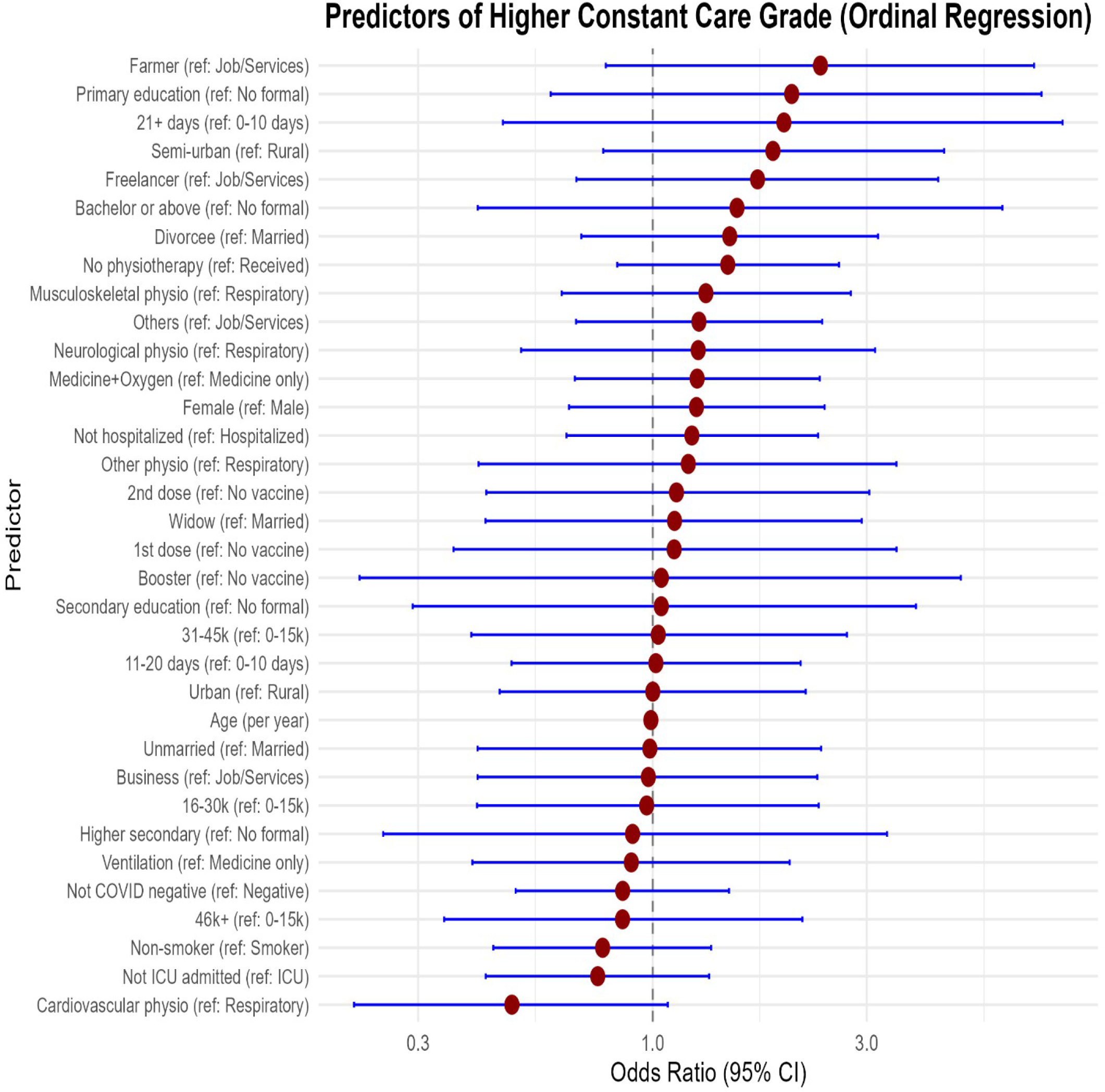
Forest plot of predictors for constant care ordinal regression. Adjusted odds ratios with 95% confidence intervals for predictors included in the ordinal logistic regression model.

**Figure 4.**
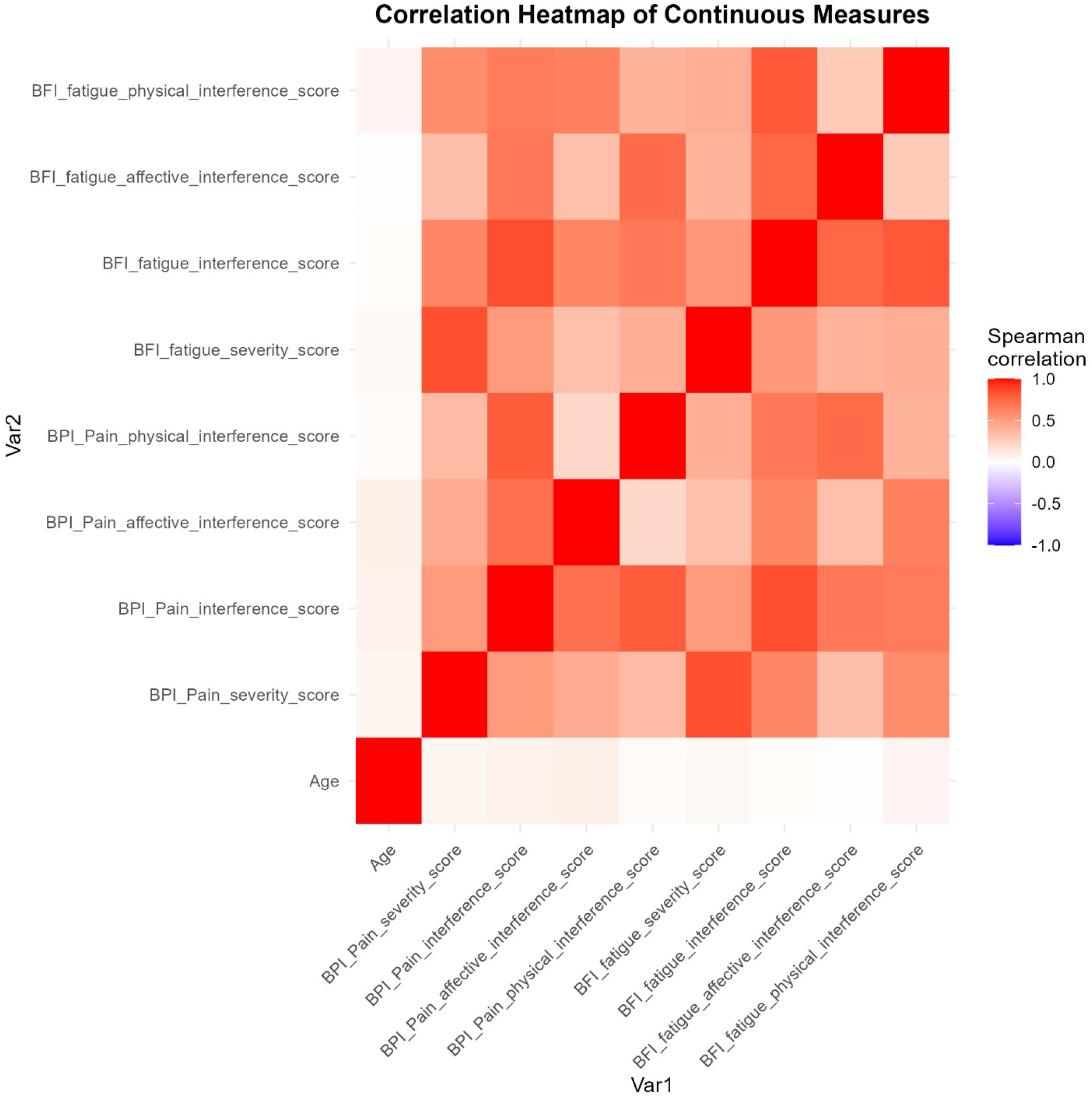
spearman correlation heatmap of continuous measures. Pairwise Spearman correlations among age and BPI/BFI measures.

## Notes

### Competing Interest Statement

The authors have declared no competing interest.

### Author Declarations

The Institutional Review Board of Bangladesh Health Professions Institute, Centre for the Rehabilitation of the Paralysed, gave ethical approval for this work (approval reference: CRP-BHPI/IRB/10/2022/673). Approval was granted at the 33rd Institutional Review Board meeting held on September 24, 2022.

