## Supplementary Statistical Tables for "Functional Impairment, Pain, and Fatigue Among Long COVID Survivors in Bangladesh: A Multisite Cross-Sectional Study"

### Supplementary Table S1. Full covariate set used in adjusted models

| Covariate | Included |
|---|---|
| Age | Yes |
| Gender | Yes |
| Marital status | Yes |
| Education | Yes |
| Occupation | Yes |
| Living area | Yes |
| Income | Yes |
| Vaccination status | Yes |
| COVID-negative status | Yes |
| Hospitalisation | Yes |
| ICU admission | Yes |
| Hospital stay duration | Yes |
| Tobacco use | Yes |
| Treatment type | Yes |
| Physiotherapy receipt | Yes |
| Type of physiotherapy | Yes |
| Physiotherapy provider | Yes |

### Supplementary Table S2. Significant functional-status associations

| Outcome | Predictor | aOR (95% CI) | Wald $\chi^2$ | p-value |
|---|---|---|---|---|
| Basic ADL | Age (per year) | 1.02 (1.00–1.05) | 3.95 | 0.047 |
| Social roles | Age (per year) | 0.97 (0.95–0.99) | 6.31 | 0.012 |
| Social roles | Unmarried (ref: married) | 0.32 (0.13–0.80) | 5.80 | 0.016 |
| Social roles | Secondary education (ref: no formal) | 0.19 (0.05–0.70) | 6.17 | 0.013 |
| Social roles | Medicine + oxygen (ref: medicine only) | 0.49 (0.25–0.95) | 4.35 | 0.037 |
| Social roles | Musculoskeletal physiotherapy (ref: respiratory) | 2.70 (1.25–5.90) | 6.31 | 0.012 |
| Social roles | Neurological physiotherapy (ref: respiratory) | 3.16 (1.26–8.07) | 5.92 | 0.015 |
| Symptom checklist | Primary education (ref: no formal) | 0.21 (0.06–0.71) | 6.31 | 0.012 |

### Supplementary Table S3. Significant pain/fatigue associations

| Outcome | Predictor | β (95% CI) | t statistic | p-value |
|---|---|---|---|---|
| BPI pain severity | Business occupation (ref: job/services) | +2.20 (0.87–3.53) | +3.34 | 0.001 |
| BPI pain severity | ICU admission: No (ref: yes) | −0.96 (−1.83 to −0.09) | −2.16 | 0.032 |
| BPI pain severity | Tobacco use: No (ref: yes) | −1.20 (−2.04 to −0.35) | −2.78 | 0.006 |
| BPI pain severity | Booster vaccine (ref: no vaccine) | −3.22 (−5.52 to −0.92) | −2.73 | 0.007 |

| Outcome | Predictor | β (95% CI) | t statistic | p-value |
|---|---|---|---|---|
| BPI pain interference | Hospitalisation: No (ref: yes) | −2.46 (−4.58 to −0.34) | −2.27 | 0.024 |
| BPI pain interference | Tobacco use: No (ref: yes) | −2.34 (−4.12 to −0.56) | −2.57 | 0.011 |
| BPI pain interference | Booster vaccine (ref: no vaccine) | −10.01 (−14.81 to −5.21) | <−3.34 | <0.001 |
| BPI pain affective interference | Occupation: Others (ref: job/services) | +1.18 (0.13–2.22) | +2.20 | 0.029 |
| BPI pain affective interference | Living area: Semi-urban (ref: rural) | +1.55 (0.06–3.04) | +2.05 | 0.042 |
| BPI pain affective interference | Physiotherapy: No (ref: received) | −1.16 (−2.11 to −0.21) | −2.41 | 0.017 |
| BPI pain affective interference | Booster vaccine (ref: no vaccine) | −4.42 (−6.93 to −1.91) | <−3.34 | <0.001 |
| BPI pain physical interference | Marital status: Widow (ref: married) | +2.52 (0.15–4.90) | +2.09 | 0.038 |
| BPI pain physical interference | Tobacco use: No (ref: yes) | −1.87 (−3.25 to −0.50) | −2.68 | 0.008 |
| BPI pain physical interference | Booster vaccine (ref: no vaccine) | −5.58 (−9.31 to −1.85) | −2.73 | 0.004 |
| BFI fatigue severity | Marital status: Unmarried (ref: married) | +1.22 (0.06–2.38) | +2.06 | 0.041 |
| BFI fatigue severity | Business occupation (ref: job/services) | +1.45 (0.33–2.57) | +2.54 | 0.012 |
| BFI fatigue severity | Physiotherapy: No (ref: received) | −0.89 (−1.62 to −0.15) | −2.37 | 0.019 |
| BFI fatigue severity | Booster vaccine (ref: no vaccine) | −4.09 (−6.04 to −2.14) | <−3.34 | <0.001 |
| BFI fatigue interference | Tobacco use: No (ref: yes) | −2.10 (−3.61 to −0.58) | −2.73 | 0.007 |
| BFI fatigue interference | Booster vaccine (ref: no vaccine) | −7.07 (−11.17 to −2.97) | <−3.34 | <0.001 |
| BFI fatigue affective interference | Tobacco use: No (ref: yes) | −0.96 (−1.71 to −0.20) | −2.51 | 0.013 |
| BFI fatigue affective interference | Booster vaccine (ref: no vaccine) | −3.33 (−5.36 to −1.29) | −3.13 | 0.002 |
| BFI fatigue physical interference | Marital status: Widow (ref: married) | +2.27 (0.20–4.35) | +2.16 | 0.032 |
| BFI fatigue physical interference | Booster vaccine (ref: no vaccine) | −3.74 (−6.99 to −0.49) | −2.26 | 0.025 |

### Supplementary Table S4. Correlation test statistics

| Variable pair | Spearman's ρ | Test statistic | p-value |
|---|---|---|---|
| BPI pain severity vs BPI pain interference | 0.85 | ρ=0.85 | <0.001 |
| BPI pain severity vs BFI fatigue severity | 0.78 | ρ=0.78 | <0.001 |
| BPI pain severity vs BFI fatigue interference | 0.82 | ρ=0.82 | <0.001 |

Note: Correlation p-values are reported as <0.001 for the highlighted strong associations. The supplied results reported Spearman correlation coefficients but did not provide exact p-values; no exact p-value beyond the reported threshold is asserted.
