## Supplementary material for "Functional Impairment, Pain, and Fatigue Among Long COVID Survivors in Bangladesh: A Multisite Cross-Sectional Study": STROBE Checklist

### STROBE Checklist – Cross-Sectional Study

| STROBE item | Content | Location |
|---|---|---|
| Title/abstract | Identifies study design and summarizes key results | Title; Abstract |
| Background/rationale | Scientific background and rationale | Introduction |
| Objectives | States study objectives | Introduction |
| Study design | Presents key design elements | Methods – Study design |
| Setting | Describes study setting and facilities | Methods – Study design and setting |
| Participants | Eligibility and sampling described | Methods – Participants and sampling |
| Variables | Outcomes and predictors described | Methods – Measures |
| Data sources/measurement | Measurement instruments described and original sources cited | Methods – Measures; References 6–8 |
| Bias | Selection, confounding, and indication bias addressed | Discussion; Limitations |
| Study size | Reports final sample size | Methods; Results |
| Quantitative variables | Describes handling of continuous and categorical variables | Statistical analysis |
| Statistical methods | Ordinal logistic regression, multiple linear regression, Wald $\chi^2$/t tests, and Spearman correlation described | Statistical analysis |
| Participants | Reports participant characteristics | Results – Table 1 |
| Descriptive data | Reports outcome distributions | Results – Table 2 |
| Main results | Reports adjusted associations with confidence intervals and test statistics | Results – Tables 3 and 4 |
| Other analyses | Reports correlation analysis with Spearman's $\rho$ | Results – Correlation analysis |
| Key results | Summarizes principal findings | Discussion |
| Limitations | Discusses design, sampling, measurement, multiplicity, and rehabilitation exposure limitations | Limitations |
| Interpretation | Balanced interpretation with caution on causality | Discussion |
| Generalisability | Discusses external validity | Limitations |
| Funding | Reports funding information | Declarations |
